# Virtual reality as a strategy for intra-operatory anxiolysis and pharmacological sparing in patients undergoing breast surgeries: the V-RAPS randomized controlled trial protocol

**DOI:** 10.1101/2024.11.21.24317660

**Authors:** Joe Zako, Nicolas Daccache, Julien Burey, Ariane Clairoux, Louis Morisson, Pascal Laferrière-Langlois

**Author notes:** Corresponding author (JZ). **Funding:** This study will be funded by Dr. Pascal Laferrière-Langlois from CR-HMR and the Department of Anesthesiology and Pain Medicine. **Competing interests:** Dr PLL declares ownership interest in private companies unrelated to this work (Divocco Medical and Divocco AI). Other authors declare no competing interests. **Data availability:** Data will be made available upon request while respecting strict confidentiality.

## Abstract

**Introduction:** Virtual reality (VR) has increasingly found applications beyond leisure and video games, extending into the field of medicine. Recent studies indicate that VR can effectively reduce anxiety and pain in pediatric patients undergoing uncomfortable medical procedures, such as burn wound care. Yet, VR use in the operating room is still rare, despite a growing trend toward regional anesthesia without general anesthesia; physicians still frequently rely on pharmacological sedation to manage procedural anxiety. By leveraging VR’s anxiolytic properties, it may be possible to decrease the need for intravenous (IV) sedation which is associated with risk of adverse events like apnea and hypoxemia and delayed recovery.

**Objectives:** This study’s main objective is to explore the impact of VR on IV sedation requirements in adult patients undergoing breast surgery under paravertebral (PV) block without general anesthesia. We hypothesize that VR immersion will reduce the need for intraoperative pharmacological sedation. Secondary objectives include assessing the tolerance of patients to the VR headset, examining the impact of the chosen VR scenario on the primary outcome, evaluating the incidence of adverse effects, measuring patient satisfaction, and analyzing the output of the Nociception Level (NOL) Index among awake surgical patients.

**Material and methods:** This single center randomized controlled trial will enroll 100 patients aged 18 or above undergoing breast surgery under PV block. Participants will be randomly allocated to the VR group or the control group; both will have access to pharmacological sedation through patient-controlled sedation (PCS). Participants in the VR group will choose between three different VR scenarios and will be allowed to switch between these scenarios during surgery. The primary outcome will be the time-adjusted and weight adjusted dose of self-administered intraoperative propofol. Secondary outcomes will include patient satisfaction, adverse events, and post-anesthesia care unit length of stay (PACU LOS).

**Ethics:** This trial has been approved by the regional ethics committee (Comité d’Éthique de la Recherche du CIUSSS de l’Est de l’Île de Montréal) on September 9^th^, 2024.

**Trial registration:** ClinicalTrials.gov (July 25^th^, 2024). Unique protocol ID: 2025-3802. Trial identification number: NCT06522711.

## 1. Introduction

### 1.1. Background and rationale

Virtual reality (VR) is a novel technology that operates through computer generated three-dimensional environments with which the user can interact. These virtual experiences are accessible on various interfaces, ranging from smartphone screens that display a VR environment to fully immersive headsets and systems that provide haptic feedback, simulating the sense of touch (1). Over the past decade, the use of VR has expanded across various medical fields. This technology has been used to train residents to perform medical procedures, to allow experienced surgeons to rehearse complex surgeries and has even been used as a substitute for exposure therapy when treating patients with specific phobias or anxiety disorders (2–5).

In anesthesiology, VR immersion has been utilized during various procedures, including orthopedic surgeries of the upper extremities, hip and knee arthroplasties under regional anesthesia, dental care, burn wound treatment, and even during anesthesia induction (6, 7). It has shown significant potential in reducing anxiety and improving patient comfort in these perioperative and periprocedural contexts, although the literature on the topic mostly pertains to pediatric populations (8, 9). Notably, research indicates that interactive, gamified VR scenarios provide significantly greater anxiolysis compared to passive scenarios, likely due to their increased distractibility (10). While a reduction of procedural anxiety has also been observed in adult populations undergoing VR immersion (11), the assessment of objective outcomes, such as procedural sedative usage, has not been thoroughly explored.

Pharmacological sedation is a crucial part of awake medical procedures, primarily aimed at providing anxiolysis and, when necessary, amnesia. However, these medications can have serious side effects, such as respiratory depression, especially when used alongside opioid analgesics (12). In fact, research suggests a dose-dependent relationship between the amount of procedural sedatives administered and the length of recovery times in the post-anesthesia care unit (PACU) (13). This highlights a critical gap in the current literature, as VR could be a low-risk adjunct or alternative to procedural sedation. This is especially relevant for breast surgeries performed only under regional anesthesia, which often require substantial amounts of sedatives.

### 1.2. Objectives and hypotheses

The primary objective of this study is to determine if intraoperative virtual reality immersion reduces the use of self-administered propofol for patients undergoing breast surgery under paravertebral (PV) block.

Our secondary objectives are the following:

- Appreciate the participants’ initial enthusiasm at the idea of using a VR headset during surgery.
- Evaluate the level of anxiety before the surgery;
- Evaluate the incidence of adverse effects such as cybersickness, nausea, bradycardia, desaturation, and hypotension;
- Evaluate the time the patient spent wearing the headset;
- Evaluate the differences in sedation requirements depending on the type of VR scenario.
- Evaluate quantities of remifentanil administered;
- Evaluate quantities of ketamine administered;
- Evaluate the requirement of post-operative care and the post-anesthesia monitoring time;
- Appreciate the ease of use of the technology, enjoyment of the first scenario chosen and overall satisfaction with the experience.

This trial also has an exploratory objective, which is to investigate the use of the Nociception Level (NOL) index in awake patients undergoing surgery, and the ability to anticipate sedation self-administration via the NOL index.

Regarding our primary objective, we hypothesize that patients undergoing breast surgery under PV block with an immersive VR experience will self-administer less propofol than the control group. We also hypothesize that interactive scenarios will further reduce the requirement for sedation, when compared to non-interactive scenarios.

### 1.3. Trial design

This is the protocol for a prospective, minimal risk randomized controlled superiority trial conducted at a single center.

## 2. Methods

This protocol follows the Standard Protocol Items: Recommendations for Interventional Trials (SPIRIT) Statement guidelines (14).

### 2.1. Participants, interventions, and outcomes

#### 2.1.1. Study setting

This trial will be conducted at Maisonneuve-Rosemont Hospital, a part of the Centre intégré universitaire de santé et de services sociaux (CIUSSS) de l’est de l’île-de-Montréal (CEMTL), located in Montreal, Quebec, Canada.

#### 2.1.2. Eligibility criteria

We will recruit consenting adult patients undergoing elective breast surgery under PV block without general anesthesia.

Patients will automatically be excluded from the study if they have any of the following:

1. Hearing or visual impairment;
2. History of epilepsy, seizure, or severe dizziness;
3. Severe mental impairment;
4. Recent eye or facial surgery or wounds;
5. Inability to use the VR hand controller;
6. Allergy to one of the protocolized drugs.

#### 2.1.3. Interventions

Prior to entering the operating room, all participants will receive an explanation on the use of intraoperative patient-controlled sedation (PCS). For those in the intervention group, a short video will introduce all three VR scenarios, after which participants will select their preferred scenario, which will then be documented.

Upon arrival in the operating room, participants will undergo standard monitoring, including non-invasive blood pressure measurement, pulse-oximetry, and continuous electrocardiography using the Dräger Infinity C700 monitor (Dräger Medical, Lübeck, Germany). Throughout the entire duration of surgery, the NOL index finger probe, which is connected to the PMD-200 monitor (Medasense Biometrics Ltd, Ramat Gan, Israel) will be applied. All intraoperative data and events will be recorded on the research computer.

The attending anesthesiologist will perform PV block before surgery according to their usual practice. Only anesthesiologists with a minimum of 10 PV blocks performed within the last year will be qualified to administer this procedure to trial participants. A standardized dose of intravenous (IV) sedation will be administered to all participants prior to realization of the PV block (0.15 mg/kg of propofol) and will be repeated if necessary. Once the block is complete, the participant is positioned supine, and the surgeon is ready for disinfection, the VR headset will be applied to those in the intervention group and their chosen scenario will begin. If at any time the patient expresses a desire to discontinue VR during the surgery, we will first offer them an alternate scenario. If they reiterate their willingness to stop, with or without having experienced the new scenario, the headset will be removed for the remainder of the surgery. In case of adverse events attributable to VR gear, such as cybersickness, discontinuation of VR immersion will be at the discretion of the anesthesiologist as well as the patient. A visual representation of the VR device used as well as the possible scenario choices can be seen in Fig. 1.

**Fig. 1.**
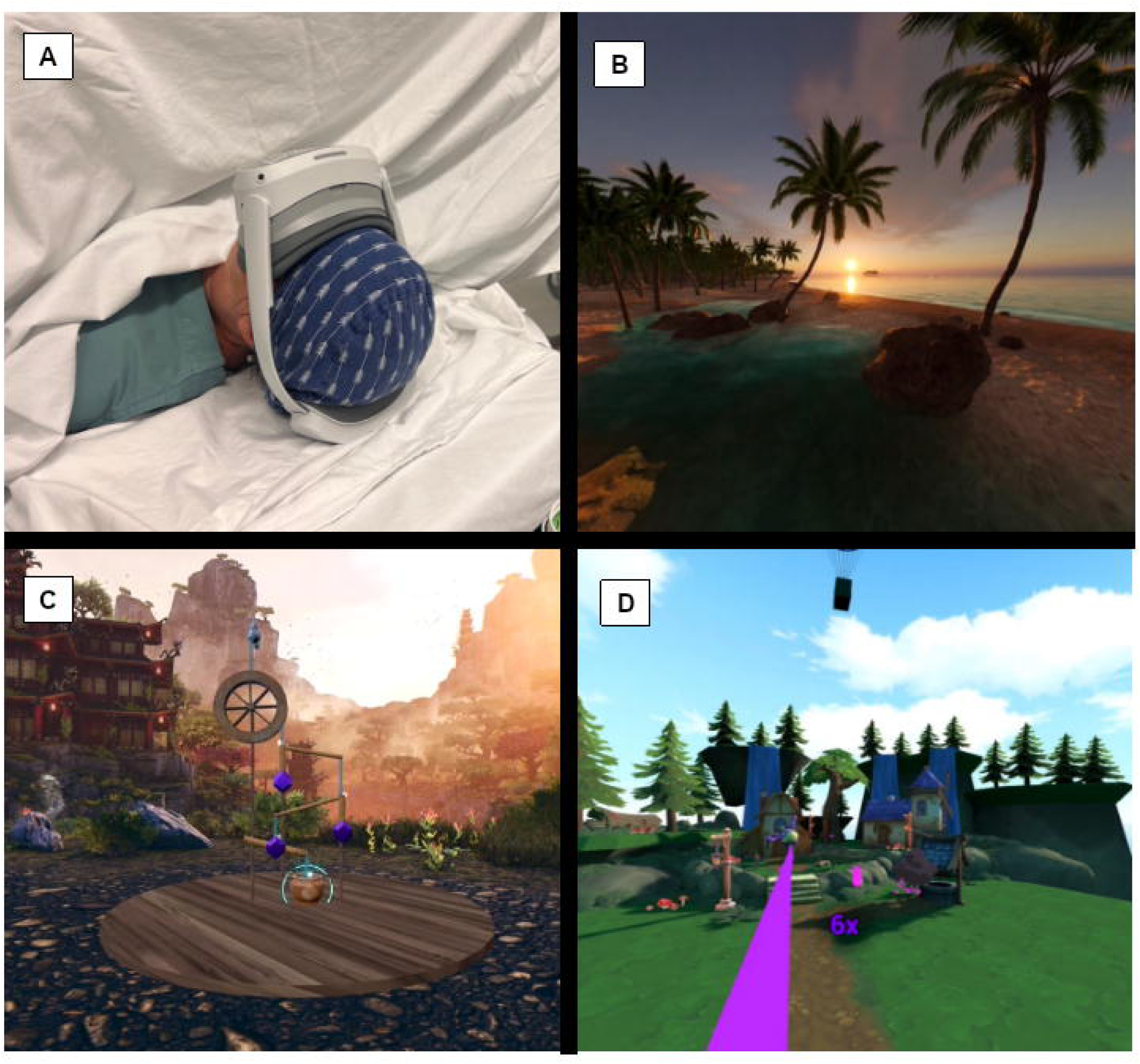
VR headset and scenario choices. Figure legend: **A** Paperplane Therapeutics VR headset; **B** Scenario 1 - Calm; **C** Scenario 2 - Puzzle; **D** Scenario 3 - Action.

In both groups, the PCS protocol will consist of 0.15 mg/kg boluses of IV propofol with a lockout period of 2 minutes; these parameters may be adjusted based on the lead anesthesiologist’s clinical judgment. For breakthrough pain, the clinician will first administer 0.5 mcg/kg of IV fentanyl and, if insufficient, 0.15 mg/kg of IV ketamine. If the desired level of analgesia and sedation is not achieved with these measures, the anesthesiologist will switch the participant to general anesthesia.

#### 2.1.4. Outcomes

The primary outcome is the time-adjusted and weight-adjusted average or median self-administered dose of propofol in mcg/kg/min.

Secondary outcomes include:

- Initial enthusiasm at the idea of using a VR headset during surgery assessed on a 10-point Likert scale for all participants, prior to intervention allocation;
- Level of anxiety before the surgery, once allocation is confirmed and known to the patient;
- Incidence of intraoperative and postoperative adverse events such as bradycardia, desaturation and hypotension as well as subjective adverse events, such as cybersickness or nausea;
- Average or median total time, in minutes, during which the VR headset was worn, as well as the percentage of that time relative to the overall duration of the surgery;
- Average or median total duration in minutes spent by the patient on the VR scenario chosen, and the order in which VR scenarios were presented;
- Percentage of patients that switched scenarios;
- Percentage of patients that removed the headset before the end of the surgery;
- Weight-adjusted average or median administration of intraoperative IV fentanyl in mcg/kg;
- Weight-adjusted average or median administration of intraoperative IV ketamine in mg/kg;
- PACU length of stay (LOS);
- Ease of use of the technology, enjoyment of the first scenario chosen and overall satisfaction with the experience assessed post-operatively on a 10-point Likert scale in the intervention group only.

Exploratory outcome:

- NOL index readings over time per patient, assessed continuously.

#### 2.1.5. Participant timeline

The research team will review the elective surgical schedule at Maisonneuve-Rosemont Hospital to identify eligible patients at least one week before their surgery. After institutional consent is obtained, patients’ medical charts will be screened based on the study’s eligibility criteria. Potential participants will be contacted by telephone to explain the project and answer supplementary questions.

One their interest in the project is confirmed, candidates will meet with the research team on the day of their surgery to address any further queries, sign the consent form, and complete the standard preoperative questionnaire. After randomization, participants will undergo regional anesthesia and surgery, with or without the VR headset, based on their allocation. A variety of preoperative, intraoperative and postoperative assessments relating to our outcomes of interest will be performed. Their involvement in the study will conclude upon discharge from the PACU. A detailed schedule of enrollment, interventions, and assessments is provided in Fig. 2.

**Fig. 2.**
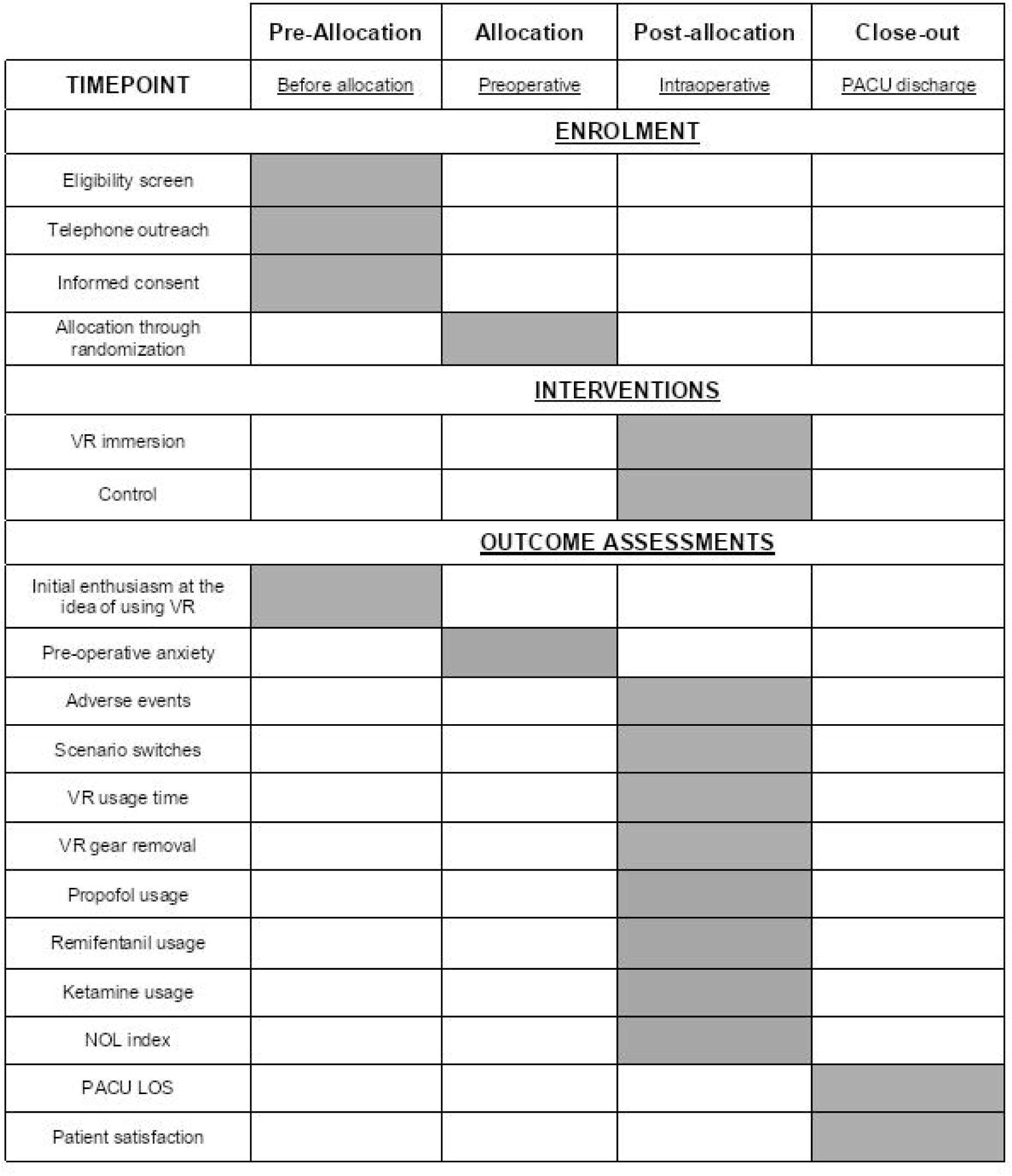
Schedule of enrolment, interventions and assessments.

#### 2.1.6. Sample size

The available literature on sedation requirements for adult patients using intraoperative VR headsets during surgeries under regional anesthesia is limited and heterogeneous, with studies reporting up to 93% reductions in sedation requirements (15), and others reporting no significant changes (16). For our sample size calculation, we estimated a 30% reduction of propofol usage in the VR group compared to the control group. With a 95% confidence level and 80% power, a total of 90 patients are required to achieve the desired precision for our estimate. To account for a potential dropout rate of 10.0%, the total sample size was increased to 100 patients.

#### 2.1.7. Recruitment

Recruitment for this trial is anticipated to begin on October 30^th^, 2024. By screening all potential candidates and contacting them by telephone beforehand, we will ensure optimal participant enrolment. We expect to complete recruitment by September 2026. Experimentation and data collection will also occur throughout this period.

### 2.2. Assignment of interventions

Electronic randomization of the participants will be performed by a statistician using a computer-generated randomized sequence with a variable block size unknown to the investigators, and then implemented in the REDCap application (Vanderbilt University) (17). Each participant’s allocation will be sealed in an opaque envelope and handed to the dedicated research staff not involved in patient care. The envelope will be opened at the patient’s entry into the operating ward, after confirming that the surgery will be performed. Due to the nature of VR, the intervention cannot be blinded to participants or personnel after allocation.

### 2.3. Data collection, management and analysis

#### 2.3.1. Data collection methods

We will collect data at baseline, intraoperatively and postoperatively to assess primary and secondary outcomes. Participant socio-demographic information, medical history and current medication, will be collected from their medical chart. Their education level and familiarity with VR technology will also be recorded through a self-reported preoperative questionnaire. Pre-operative anxiety levels will be assessed using the Amsterdam Preoperative Anxiety and Information Scale (APAIS).

NOL index data from the PMD-200 medical monitor will be automatically collected by the device. Other intraoperative events, such as propofol self-administration, as well as the use of remifentanil or ketamine, will be manually recorded. Postoperative outcomes will be collected through questionnaires administered to participants prior to their discharge from the PACU, at which point we will also record PACU length of stay (LOS). Adverse events of any kind will be documented when spontaneously reported or observed clinically.

We will monitor and document the number of participants who withdraw from the study. If a participant wishes to discontinue their involvement, they will be removed from the study at their request, and no further data will be collected or retained after their withdrawal.

#### 2.3.2. Data management

Data from this study will be collected and managed using REDCap electronic data capture tools hosted at Maisonneuve-Rosemont hospital. Research staff will systematically double-check all data entries for completeness and accuracy before final submission. Each participant will be assigned a unique identification number upon enrollment to maintain confidentiality and ensure anonymization.

All collected data will be securely stored on a designated research computer, which will remain offline and be located in a restricted area in Maisonneuve-Rosemont hospital. Notably, this computer will utilize the integrated event-tagging system of the PMD-200 or the BetterCare software (provided by Dräger, Lübeck, Germany), which is connected to the Dräger anesthesia workstation. Usage of this computer will be limited to authorized research personnel, who will need encrypted logins to access study data.

To promote data quality, double data entry will be implemented for our primary outcome. Furthermore, regular, informal audits will be conducted by the research team to verify the accuracy of the study data.

After study completion, electronic data will be retained on secure, encrypted servers for a period of 7 years, as per local institutional and ethical guidelines. Physical copies of patient information, such as consent forms, will be securely stored in a locked file cabinet in the anesthesiology department.

#### 2.3.3. Statistical methods

Descriptive statistics will be used to summarize the data by group. Primary outcome and secondary continuous parameters will be presented as means with standard deviations, or medians with interquartile ranges if the data is skewed or non-normally distributed. Categorical variables will be reported as frequencies (%). 95% confidence intervals (CIs) for proportions or mean/median differences will be presented based on the type of endpoint analyzed, and statistical significance will be determined using an alpha level of 0.05. Parametric, two-sample t-tests will be performed for normally distributed outcomes. Otherwise, a non-parametric Wilcoxon test will be performed.

Univariate analyses will be performed to explore relationships between the primary outcome and other potential variables. Logistic regression will be used to analyze each independent variable’s association with the primary outcome. If relevant, multivariate analyses will be conducted to assess the influence of additional independent variables. Subgroup analyses will also be performed to explore how various factors, such as the selected VR scenario, may impact the primary and secondary outcomes to address our secondary objectives. In the case of conversion to general anesthesia, the participant will be considered a protocol breach for the purposes of the final statistical analysis.

As part of our exploratory analysis, we will investigate the relationship between the NOL index and propofol self-administration. Specifically, we will discretize NOL index data by qualifying what we consider a spike (e.g. any score above a threshold of 50) and assess whether these spikes can predict propofol self-administration timepoints.

All statistical analyses will be conducted using SAS, SPSS, or Python programming language with VS Code or Jupyter Notebook software.

### 2.4. Monitoring

#### 2.4.1. Data monitoring

The study is considered low-risk and will not require a formal data monitoring committee. The principal investigator (PI) or designated personnel will regularly review the data on a monthly or bi-weekly basis to assess study completeness, enrollment progress, protocol deviations, participant dropouts, and adverse events. The study will also be continuously overseen by the regional ethics committee.

#### 2.4.2. Potential harms

The devices used in the study include a VR headset wirelessly paired with a tablet for broadcasting immersive scenarios. These devices do not interfere with intraoperative monitoring, and the risk of adverse events associated with their use is very low.

Any incidence of adverse effects, spontaneously reported or directly observed, will be documented and assessed as part of our secondary outcomes. Participants at higher risk of serious side effects will be pre-emptively screened and excluded from the study.

#### 2.4.3. Auditing

There is no formal plan for auditing or inspection in this study.

## 3. Ethics and dissemination

### 3.1. Research ethics approval

This protocol has been approved by the regional ethics committee (Comité d’éthique en recherche du CIUSSS de l’Est de l’Île de Montréal) on September 9^th^, 2024.

### 3.2. Protocol amendments

Any changes to the study protocol will be submitted for review by the regional ethics committee and updated on ClinicalTrials.gov. These amendments will be communicated to participants and study personnel where necessary and will be documented in the final study report. In the event of study termination, participants will be notified, and data will be handled according to ethical guidelines.

### 3.3. Consent or assent

During recruitment, potential participants will be contacted by the research team via phone to explain the project. The communication script used during these calls has been approved by the regional ethics committee. Prior to surgery, the research team will meet with candidates in person to answer any remaining questions. If participants wish to proceed, informed consent will be obtained and signed. The form used to obtain written consent by the research staff will be made available upon request to the corresponding author.

### 3.4. Confidentiality

Protected health information will not be re-used or disclosed to third parties except as required by law, for authorized oversight of the research, or as permitted by patient authorization. All digital and physical patient information will be stored in secure environments (see 2.3.2. Data management). All team members involved in the study will receive proper training and adhere to confidentiality protocols to protect participant privacy. The study will fully comply with all applicable laws, regulations, and guidelines.

### 3.5. Declaration of interests

PLL declares ownership in private companies unrelated to this work (Divocco Medical and Divocco AI). All other authors declare no competing interests.

### 3.6. Access to data

Only the principal investigator (PI) and co-investigators will have access to the final trial dataset. Access to de-identified participant-level data can be requested from the corresponding author, while ensuring strict confidentiality.

### 3.7. Post-trial care

Ensuring subject safety is a top priority for the research team and hospital staff. In the unlikely chance of an unexpected serious adverse event, the PI, Dr. Pascal Laferrière-Langlois, will be immediately notified and prompt actions will then be taken to provide appropriate care.

### 3.8. Dissemination policy

We aim to publish the trial results in a mid-impact factor journal in the field of anesthesia or medical technology. Patients and the public were not involved in the design, conduct, or reporting of this research, and will not be involved in its dissemination. Only those that contribute significantly to the advancement and publication of the study will be considered eligible for authorship.

## 4. Discussion

### 4.1. Limitations of study design

#### 4.1.1. Unblinded personnel

Due to the nature of VR interventions, neither the patient, the anesthesiologist or surgeon can be blinded to group allocation. However, using PCS minimizes the anesthesiologist’s influence on propofol administration, reducing performance bias.

#### 4.1.2. Single-center study

This trial will be conducted at a single hospital, which may limit the generalizability of results. However, the expected sample size is larger than many similar studies, which should improve the robustness and reliability of the findings.

#### 4.1.3. Short-term outcomes

The study focuses solely on short-term outcomes, concluding once participants are discharged from the PACU. While long-term outcomes like post-operative delirium or cognitive impairment were not considered essential for this research, future studies could explore these longer-term effects.

## 5. Author contributions

Conceptualization – JZ, ND, JB, LM, PLL; Investigation – JZ, JB and PLL; Methodology – JZ, ND, JB, AC, LM and PLL; Supervision – PLL; Writing (Original Draft Preparation) – JZ and JB; Writing (Review & Editing) – ND, LM and PLL.

All authors have read and approved the final manuscript.

## Data Availability

No datasets were generated or analysed during the current study. All relevant data from this study will be made available upon study completion.

## 6. Acknowledgements

We thank Paperplane Therapeutics for providing the VR hardware and software. We also thank the whole research team at the Laboratory of Innovative Anesthesia in Montreal (LIAM) for their assistance in research organization.

## Appendix A World Health Organization trial registration dataset

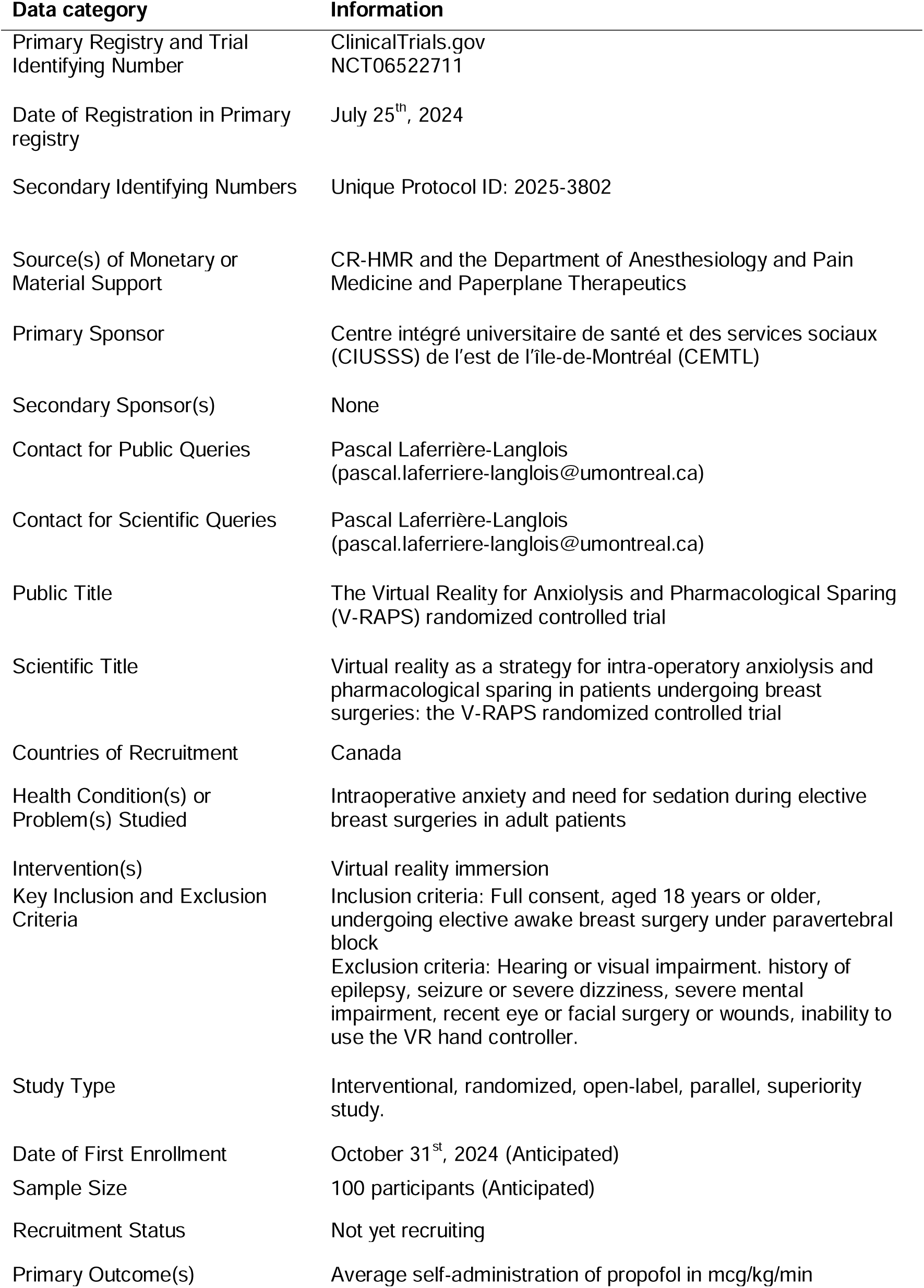
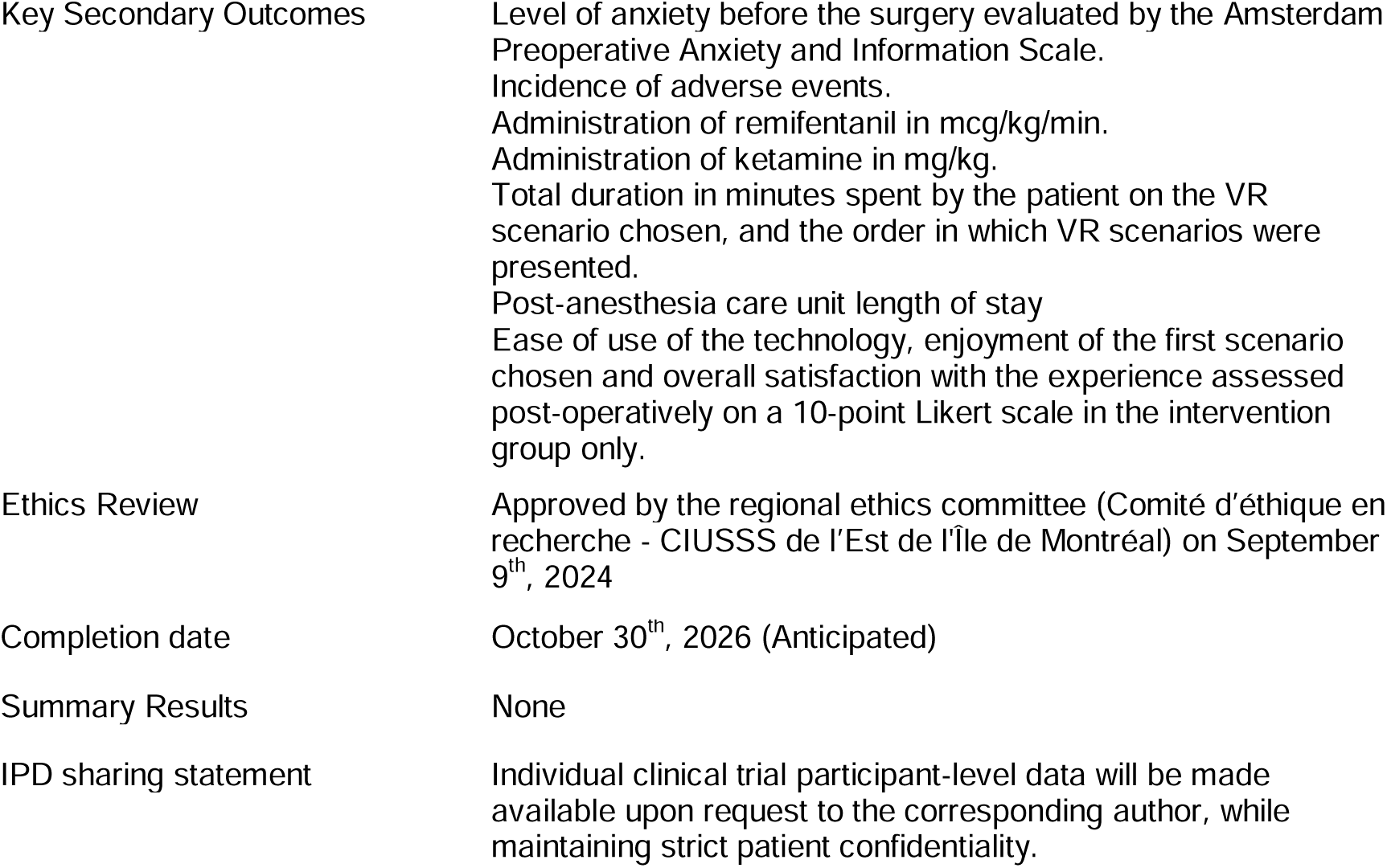

## Notes

### Funding Statement

The author(s) received no specific funding for this work.

### Author Declarations

The regional ethics committee for the Integrated University Center for Health and Social Services of the east of the island of Montreal gave ethical approval for this work on September 9th, 2024.

## References

1. Boutin J, Kamoonpuri J, Faieghi R, Chung J, de Ribaupierre S, Eagleson R. Smart haptic gloves for virtual reality surgery simulation: a pilot study on external ventricular drain training. Front Robot AI. 2023;10:1273631.

2. Carl E, Stein AT, Levihn-Coon A, Pogue JR, Rothbaum B, Emmelkamp P, et al. Virtual reality exposure therapy for anxiety and related disorders: A meta-analysis of randomized controlled trials. J Anxiety Disord. 2019;61:27–36.

3. Chumnanvej S, Chumnanvej S, Tripathi S. Assessing the benefits of digital twins in neurosurgery: a systematic review. Neurosurg Rev. 2024;47(1):52.

4. Kuhn AW, Yu JK, Gerull KM, Silverman RM, Aleem AW. Virtual Reality and Surgical Simulation Training for Orthopaedic Surgery Residents: A Qualitative Assessment of Trainee Perspectives. JB JS Open Access. 2024;9(1).

5. Yi WS, Rouhi AD, Duffy CC, Ghanem YK, Williams NN, Dumon KR. A Systematic Review of Immersive Virtual Reality for Nontechnical Skills Training in Surgery. J Surg Educ. 2024;81(1):25–36.

6. Boyce L, Jordan C, Egan T, Sivaprakasam R. Can virtual reality enhance the patient experience during awake invasive procedures? A systematic review of randomized controlled trials. Pain. 2024;165(4):741–52.

7. Hitching R, Hoffman HG, Garcia-Palacios A, Adamson MM, Madrigal E, Alhalabi W, et al. The Emerging Role of Virtual Reality as an Adjunct to Procedural Sedation and Anesthesia: A Narrative Review. J Clin Med. 2023;12(3).

8. Eijlers R, Utens E, Staals LM, de Nijs PFA, Berghmans JM, Wijnen RMH, et al. Systematic Review and Meta-analysis of Virtual Reality in Pediatrics: Effects on Pain and Anxiety. Anesth Analg. 2019;129(5):1344–53.

9. Wang Y, Guo L, Xiong X. Effects of Virtual Reality-Based Distraction of Pain, Fear, and Anxiety During Needle-Related Procedures in Children and Adolescents. Front Psychol. 2022;13:842847.

10. Yamashita Y, Aijima R, Danjo A. Clinical effects of different virtual reality presentation content on anxiety and pain: a randomized controlled trial. Sci Rep. 2023;13(1):20487.

11. Kodvavi MS, Asghar MA, Ghaffar RA, Nadeem I, Bhimani S, Kumari V, et al. Effectiveness of virtual reality in managing pain and anxiety in adults during periprocedural period: a systematic review and meta-analysis. Langenbecks Arch Surg. 2023;408(1):9.

12. Nordt SP, Clark RF. Midazolam: a review of therapeutic uses and toxicity. J Emerg Med. 1997;15(3):357–65.

13. Ma H, Wachtendorf LJ, Santer P, Schaefer MS, Friedrich S, Nabel S, et al. The effect of intraoperative dexmedetomidine administration on length of stay in the post-anesthesia care unit in ambulatory surgery: A hospital registry study. J Clin Anesth. 2021;72:110284.

14. Chan AW, Tetzlaff JM, Gotzsche PC, Altman DG, Mann H, Berlin JA, et al. SPIRIT 2013 explanation and elaboration: guidance for protocols of clinical trials. BMJ. 2013;346:e7586.

15. Faruki AA, Nguyen TB, Gasangwa DV, Levy N, Proeschel S, Yu J, et al. Virtual reality immersion compared to monitored anesthesia care for hand surgery: A randomized controlled trial. PLoS One. 2022;17(9):e0272030.

16. Huang MY, Scharf S, Chan PY. Effects of immersive virtual reality therapy on intravenous patient-controlled sedation during orthopaedic surgery under regional anesthesia: A randomized controlled trial. PLoS One. 2020;15(2):e0229320.

17. Harris PA, Taylor R, Thielke R, Payne J, Gonzalez N, Conde JG. Research electronic data capture (REDCap)--a metadata-driven methodology and workflow process for providing translational research informatics support. J Biomed Inform. 2009;42(2):377–81.

